# A mixed methods analysis of youth mental health intervention feasibility and acceptability in a North American city: perspectives from Seattle, Washington

**DOI:** 10.1101/2023.06.23.23291816

**Authors:** Augustina Mensa-Kwao, Ingrid Sub Cuc, Tessa Concepcion, Christopher G. Kemp, Matthew Hughsam, Moitreyee Sinha, Pamela Y. Collins

**Author notes:** Corresponding author: Pamela Y. Collins.

## Abstract

**Introduction:** In March 2021, the Governor of Washington declared a youth mental health crisis. State data revealed high rates of youth suicide and inadequate access to services. This mixed-methods study examines youth and adult perspectives on mental health service gaps and opportunities in Seattle by assessing needs, feasibility, and acceptability of interventions to support youth mental health.

**Methods:** We interviewed 15 key informants to identify the contextual, structural, and individual-level factors that increase the risk of poor mental health and deter access to care among young people. We complimented these data with a cross-sectional 25-item survey of 117 participants in King County to assess the feasibility and acceptability of interventions for youth mental health. We conducted a deductive thematic qualitative analysis of the interviews and performed descriptive analyses of the quantitative data, using t-tests and χ^2^ tests to summarize and compare participant characteristics stratified by age group.

**Results:** Qualitative informants attributed challenges to youth mental health to social and relational problems. Example interventions included creating environments that increase belonging and implementation of culturally congruent mental health services. Quantitative study participants rated all evidence-based mental health interventions presented as highly acceptable. However, youth preferred interventions promoting social connectedness, peer support, and holistic approaches to care, while non-youth preferred interventions focused on suicide, alcohol, and substance abuse prevention. Both key informants and survey participants identified schools as the highest priority setting for mental health interventions. There were no significant differences among quantitative outcomes.

**Conclusion:** Our findings highlight the need for reducing social isolation and increasing social connectedness to support youth mental health. Schools and digital tools were preferred platforms for implementation. Engaging multiple stakeholders, especially young people, and addressing cultural needs and accessibility of mental health resources are important pre-implementation activities for youth mental health intervention in a US city.

## Introduction

Psychosocial stressors induced by the COVID-19 pandemic have precipitated a mental health crisis for young people in cities around the United States. The U.S. Surgeon General issued an advisory on youth mental health in late 2021, signaling the public health significance of the issue and the need for “the nation’s immediate awareness and action (1).” In Seattle, Washington, hospitals and emergency rooms endured unprecedented rates of admission for psychiatric complaints throughout 2021 (2). The state’s Governor declared a youth mental health crisis in March of 2021, marking the need to prioritize youth well-being (2). However, challenges to youth mental health were growing well before the pandemic.

In Washington State, the youth suicide rate stood at 11.4 per 100,000 in 2016; however, by 2020, it had risen to 15.7 per 100,000 (3). Currently, approximately 22% of youth aged 15-24 in the state of Washington are estimated to live with a mental disorder, while 12% are estimated to live with a substance use disorder (4). However, access to standard mental health services is limited in many parts of the state, given workforce shortages (5).

Seattle’s social context, notable for rising rates of gun violence, racial and ethnic disparities in the criminal justice system and in health outcomes, as well as housing instability and homelessness, significantly shapes the mental health trajectories of young people (6–8). Recent policies and programs seek to support the social and mental wellbeing of youth and families. Best Starts for Kids (BSK), an initiative launched in 2015, invests in early child development, and youth and family initiatives throughout King County (9). Seattle is also known for community activism and sustainable urbanization that supports community well-being and works to reduce social risk factors that can contribute to poor mental health outcomes (10).

These efforts matter. While urban environments can pose high risks for poor mental health (11–13), they can also offer mental health-promoting environments, institutions, and services that are accessible, affordable, and culturally appropriate to diverse populations, supporting youth mental health through promotion, prevention and care (13–16). Few cities adopt an intersectoral approach to address the mental health needs of adolescents and young adults, taking advantage of resources within and outside of the health system. In response to these needs, the citiesRISE consortium, a global platform committed to transforming the state of mental health policy and practice in cities around the world (17), enables young people to drive action in their communities by positioning them as expert partners with local stakeholders and by creating a unified youth voice. citiesRISE selected Seattle as a primary site to initiate engagement around youth mental health.

In 2019, a citiesRISE landscaping assessment of youth mental health identified opportunities to engage and support young people in Seattle. These opportunities included promoting racial equity, expanding the definition of mental health, and addressing substance abuse, workforce development, and peer support among youth. The same year, we hosted a multistakeholder roundtable discussion at the University of Washington to identify priorities and opportunities for supporting youth mental health in Seattle. The current study aims to ascertain in greater detail the kinds of support across the mental health care continuum recommended by young people and key stakeholders who could assist with implementation. We examine, quantitatively and qualitatively, the feasibility, acceptability of specific mental health interventions for youth in Seattle.

## Methods

### Design

We used a mixed methods study design to survey public policy informants, young people, and adults in Seattle/King County a) on the contextual, structural, and individual level factors that place young people at risk for poor mental health and deter access to care, b) to identify opportunities for interventions, and c) to determine feasibility and acceptability of interventions that could support youth mental health.

We employed these methods:

1. Qualitative key-informant interviews with 15 research participants representing youth and public policymakers and
2. A cross-sectional, quantitative anonymous survey, distributed electronically, of 117 young people and youth allies in the Seattle region with the option for open-ended feedback.

The research study received approval from the University of Washington Human Subjects protocols STUDY00007988 and STUDY00008904. Qualitative research participants received a study information sheet and provided verbal informed consent (adults) or assent without parental consent (participants under age 18). Verbal consent was documented on audio recordings of adults and with gift card payment of youth informants. Consent for quantitative survey participants was embedded in the online survey; participants under the age of eighteen assented without parental consent. The UW research team had access to data with identifiers for the qualitative sample and the quantitative study participants.

### Qualitative study design and sample

We interviewed participants with knowledge of mental health employed in public service (city and county government) and young people (17 years and older) living in King County by recruiting a convenience sample of 15 key informants between September and December 2019. We recruited five adult key-informants in public service during a series of meetings to orient local policy makers and mental health experts to citiesRISE activities and aims. We used snow-ball sampling through youth networks known to the research team members to recruit youth key-informants from two different groups: 1) young people engaged in youth-focused initiatives in Seattle (N= 4) and 2) young people working or studying in the Seattle community (N=6).

To ensure consistency, we conducted all interviews in English using a standardized interview guide that posed similar questions to each participant. The topics covered mental health needs among young people, common mental health problems, gaps in mental health services, opportunities for intervention, and opportunities for youth engagement. Youth participants received a $50 gift card for their participation. Participants completed one interview. All interviews were recorded and transcribed for analysis.

### Qualitative data analysis

We conducted a thematic analysis of the interview data driven by our research question: what are the gaps and opportunities for youth mental health interventions in Seattle/King County? After reading the transcripts multiple times, we created case summaries and conducted an in-depth indexing of the interviews, organizing them according to the primary themes of the interview guide. We used this guide to develop a deductive code list. The research team met several times to conduct open coding, review data coded, introduce new codes, and to resolve discrepancies in coding. We created a codebook based on these discussions and coded the remaining interviews. The current paper presents findings on mental health risks and needs, gaps in care, intervention opportunities, and youth engagement.

### Quantitative study design and sample

#### Sample and Data Collection

We invited young people and adults in the Seattle/King County area to complete brief, structured, online questionnaires from February through March 2020. Questionnaires took approximately fifteen minutes to be complete and were available in English. Participants were recruited through fliers and links that were disseminated through school- and community organization-based mailing lists and through an Instagram promotion targeting young people in the Seattle/King County region aged 17-24. Adults over the age of 24 were also eligible to participate. Informed by output from the roundtable discussion and preliminary qualitative data findings, we designed a twenty-five-item questionnaire. We assessed participants’ sociodemographic characteristics (i.e., gender, race/ethnicity, age, education, and employment), priorities and unmet needs related to mental health, barriers to mental health, and preferences for novel services and interventions for mental health (S1 Appendix). An error in the REDCap survey meant that data on age in years are missing for n=65 participants, though all participants self-reported being over the age of 16, and all self-identified as “youth” or “non-youth.”

#### Measures

Demographic information included participant age (in years), gender identity (woman, man, nonbinary), race or ethnicity (Alaska Native, American Indian/Native American, Asian, Asian – South Asian, Asian – Southeast Asian, Black/African American, Black African, Hispanic/Latinx/Chicanx, Middle Eastern, Native Hawaiian, Pacific Islander, White/European American, as well as any racial/ethnic identity not listed), youth identity status (youth or non-youth), birth year, employment status (studying, employed, self-employed/freelance, interning, part-time, unemployed-looking for work, unemployed-not looking for work, homemakers, military, retired, not able to work, or other), education level (no formal education, pre-school to 8th grade, some high school with no diploma, high school graduate with a diploma or equivalent (e.g., GED), some college credit with no degree, trade/technical/vocational training, associate degree, Bachelor’s degree, Master’s degree, Professional degree, Doctorate degree), South Seattle residence (yes or no), and zip code.

#### Mental Health Priority Issues and Satisfaction

Relative priority of nine different mental health-related issues (overall mental health, alcohol and substance use, depression, anxiety and stress, schizophrenia and severe mental illness, suicide and self-harm, homelessness, poverty, racism and inequality, and social connectedness) was assessed with a three-level Likert scale, where each issue was rated as not important, somewhat important, or very important. Satisfaction with these same nine mental health-related issues was assessed with a three-level Likert scale, where respondents rated their perception of efforts to address each issue as not satisfied, somewhat satisfied, or very satisfied.

#### Barriers to Mental Health

Participants were presented with a list of potential barriers to youth mental health and asked to choose the three most important barriers. The barriers included lacklack of support from peers, lack of support from parents/guardians and family, lack of future economic opportunity (e.g., jobs), lack of future educational opportunity (e.g., college), lack of after-school activities, lack of support from the school system, lack of access to quality health care, lack of awareness and skills related to mental health, exposure to violence, exposure to racism and social injustice, unstable or unavailable housing (housing insecurity), and other barriers not listed above as identified by participants.

#### Mental Health Intervention Platforms

Preferences for mental health intervention platforms and approaches to improve youth mental health were assessed by asking respondents to select two ideal platforms and two priority services. The following mental health intervention platforms were presented: schools, community centers, churches, online platforms, workplaces, clinics and other health care settings, in public/on the street, or other places not listed above as identified by participants. Participants selected from the following priority services: positive youth-led mental health messaging developed by youth for youth, training in resilience and self-care, training in awareness and peer support, provision of safe spaces, access to counseling and treatment aligned with the values and traditions of the community, housing and social services, employment opportunities and career counseling, or other services, programs, or activities not listed above as identified by participants.

#### Acceptability of Intervention Measure (AIM)

Four specific evidence-based interventions or services were described in detail, and acceptability of each intervention or service was assessed using the 4-item Acceptability of Intervention Measure (AIM) (18).These interventions included the Friendship Bench psychosocial support model, the provision of safe spaces, peer navigation and support services, and arts-based mental health promotion and suicide prevention in schools. Items assess level of approval (e.g., “Friendship Bench would meet my approval”), appeal (e.g., “Friendship Bench would be appealing to me”), liking (e.g., “I would like Friendship Bench”), and willingness to welcome (e.g., “I would welcome Friendship Bench”) the interventions. Participants rated their level of agreement or disagreement with each statement using a 5-point Likert scale, ranging from “completely disagree” to “ completely agree.” The scale provides a numerical rating that reflects the participant’s perception of the intervention’s acceptability.

### Quantitative Data Analysis

Descriptive analyses, including t tests and χ2 tests, were conducted to summarize and compare participant characteristics stratified by age group (i.e., self-reported youth vs non-youth). Proportions were calculated for categorical responses. AIM summary scores were created by averaging respective item responses and compared across age strata using t tests. In addition to the age-related data missingness identified above, there was a moderate amount of missingness due to incomplete survey responses by participants. Given that only descriptive statistical approaches were used, complete case analysis was deemed appropriate. All analyses were performed in Stata and RStudio 2022.07.2 (19, 20).

## Results

### Context

Seattle, the 22^nd^ largest city in the United States, is notable for rapid population growth over the past decade, largely attributed to an expanding workforce required by technology companies in the region. These factors yielded an increasing median household income (21), gentrification, and a growing epidemic of homelessness. The urban landscape continues to shift as developers design higher density communities and as lower income residents move southward and northward to escape the rising cost of living. In the context of these rapid social changes, concerns about youth mental health in the region have intensified in recent years.

The data collection for the current study was underway when the University of Washington (UW) Global Mental Health Program, citiesRISE, and the UW Population Health Initiative hosted a roundtable discussion of factors in the city that affect adolescent mental health and wellbeing. The participants were 38 invitees from UW (students, faculty, staff), local non-governmental organizations, and city government. They identified action steps for youth mental health intervention (**Table 1),** some of which could inform the implementation of individual level interventions.

**S1 Table:** Actions and innovations for an adolescent mental health-friendly Seattle.

| Interpersonal/Family | Community/Organizational/Policy | Environment and Society |
| --- | --- | --- |
| <u>YOUTH LEADERSHIP AND PEER SUPPORT</u><br>• Ensure process justice in designing any interventions (i.e., youth leadership, community based designs)<br>• Peer coaching for mental health, and potentially reimburse this coaching<br>• Peer groups for parents and teens (“PEPS” for teens)/peer and near peer mentoring<br>• Empowering youth<br>• Youth breakfasts for all: encouraging convening, connection, sustenance, community<br>• Empower teens: ask teens what safe/inclusive spaces look like<br>• Subsidized positions for youth in government agencies and in media<br>• Youth participatory budgeting | <u>INVESTMENT IN SERVICES and INCENTIVES</u><br>• Increase wages for front-line workers<br>• Reimburse clinicians for Evidence Based Practices at higher rates<br>• Increase wraparound services<br>• Incorporate Evidence Based Practices into licensure requirements<br>• Increase investment in human welfare<br>• Increase services for mental health promotion and mental health care provision<br>• Provide a “golden credit card” to give access to non-clinical holistic care and treatment for mental health conditions<br>• Establish flexible, 24 hour, full-time support systems<br>• Give youth autonomy to choose own services<br>• Give youth ability to access resources for their entire family<br>• Provide universal 100% mental health coverage for adolescents<br>• Build out mental health resources in spaces where people spend time<br><u>SCHOOLS AND EDUCATION</u><br>• Launch a campaign for mental health<br>• Educate caregivers and the public about mental health<br>• Teach emotional literacy: start early, and maintain consistent social-emotional learning from K-12 and university<br>• Create demand for socioemotional learning among parents<br>• Teach history, health, change coursework, include inclusive materials/topics<br>• Establish restorative justice practices in all systems<br>• Use Individualized Education Program (IEP)/504s/school accommodations<br>• Learn from other cultures for successful models<br>• Train first responders to manage mental health crises vs causing more trauma, learn de-escalation<br><u>TECHNOLOGY</u><br>• Create safe social networking spaces | <u>BUILT AND NATURAL ENVIRONMENT</u><br>• Create systems and spaces of belonging<br>• In-person microcommunities<br>• Establish safe spaces for teens to “hang out”/environments to connect<br>• Establish safe spaces for consumers of mental health services<br>• Cluster youth spaces at transit hubs<br>• Make transit free of cost |

Most discussion groups raised the following threats to youth mental health in Seattle: lack of connectedness and community spaces, discrimination and bias, cost of living pressures, and lack of integrated services around mental health. Ideas for actions that could improve youth mental health in Seattle were further discussed in several groups. Youth leadership was considered to be central in devising any actions around adolescent mental health. Participants viewed peer support and mentoring as impactful ways to support youth mental health. Likewise, the groups identified families as having great potential to support youth, while recognizing that parents require their own support systems and resources in order to be advocates for youth. Additionally, group members noted that youth need environments where their emotional development is viewed as important for success. Our qualitative interviews explored some of these themes in depth.

### Qualitative interviews

We conducted qualitative interviews with 15 key informants: 5 public service informants and 10 young people or youth allies. Among the public service informants, all served in city or county government roles. Youth participants ranged in age from 17 - 31, with 7 participants under the age of 25. The group included 3 men and 7 women and represented young people with multiple cultural identities.

#### Perceived risks for poor mental health among youth in Seattle

Public service participants identified homelessness, domestic violence, access to weapons, risky home environments, substance abuse, harm by the criminal justice system, suicide, and isolation or lack of connectedness as drivers and consequences of poor mental health among youth in Seattle and King County. Youth participants and allies identified depression and anxiety, gentrification, school competition, intergenerational challenges, affordable housing, and exposure to adverse childhood events among the areas influencing youth mental health. All participants highlighted the challenges of isolation and connectedness as important factors.

##### Fighting hard to stay connected

Public service and youth/ally participants identified lack of connectedness as the most commonly reported risk factor for poor mental health among young people. A participant noted that while loneliness is often recognized as a health problem for older adults, young people also experience the “significant problem of isolation.” Gentrification was proposed as one explanation. Several informants explained that the rapid pace of gentrification led to the displacement of youth or their peers from communities where they had longstanding social connections. According to one participant, “The kids you start off with in grade school are not going to be the same kids that you end up with in high school,” as had been typical in the past. “Part of that has to do with the affordability issue of being able to live in the city over the long term,” another participant described. Participants highlighted the devastating impact of gentrification on communities—people were “really fighting so hard to…stay connected to the schools and communities they feel they have the most connection with.” As a result of this displacement, families were often scattered across the southern part of the county, disrupting informal support systems and exacerbating physical and emotional isolation.

Young people also attributed social isolation and lack of connectedness to the “Seattle freeze,” a cultural trait marked by social reserve and a lack of engagement with others. Youth participants from other regions of the country recounted the difficulty of making new friends and bewilderment when their usual gestures of friendliness were not returned. One participant confessed, “I didn’t understand why people were so distant…I remember I would like to smile at people on the street and they would look confused…that kind of isolation is a detriment to mental health and for youth for sure.”

Another youth participant linked isolation with self-blame, which reinforced young people’s social withdrawal. “Folks are like, I’m the only person who’s struggling with this and…I’m just doing it by myself…no one would…ever understand what I’m struggling with, you know.” Public service participants, who alluded to recent increases in suicides among youth in Seattle, linked risk with social isolation.

##### Mental health is taboo

Given the taboo surrounding mental health and mental illness, young people were often reluctant to discuss their symptoms, which further exacerbated their feelings of isolation. A participant noted, “There weren’t a lot of people who are open to speaking about mental health, and I think that’s also another barrier that leads people to closing everything in, not being able to speak about it and realizing…like, there’s something wrong with me, but I don’t know what it is.” Intergenerational differences in cultural understanding of mental health problems could elicit similar feelings of isolation and increased stress. In some cases, parents who had immigrated to Seattle and sought to establish a new life tended to prioritize “survival mode” over mental health, leading to a perception that mental health concerns were less important and not to be raised as a problem. Participants recognized consequences of this dismissal. A student commented, “If at home you can’t express your emotions and at school you need to study, study, work and work…then your emotions never come out…and they’re building, they’re building, they’re building, and it turns into depression…”

### Gaps in responses to mental health needs

#### No place to go

“Not having a place to go” and “not reaching out” complicated young people’s ability to access care, but the sense that services were siloed and that few options existed outside of highly structured inpatient environments was viewed as problematic by some respondents. “There is nothing really in between--a supportive program where they can, you know, relapse. If you don’t succeed, you’re failing.” A public service participant noted that the mental health system’s emphasis on numbers of beds and residential treatment conflicted with the aim to divert youth from more restrictive environments. “We’re striving to find opportunities to divert youth from those places and to be able to seek help in communities.” When youth accessed outpatient formal mental health services, the challenges of payment and confidentiality within the family arose. Some described the difficulty of not telling parents about seeking care, yet needing access to parental insurance or other financial support. Some noted that few responses outside of the health system existed and that youth would benefit from more holistic approaches that “go beyond the individual” and that were not part of formal healthcare services.

Outside of formal services, several participants noted a lack of designated spaces for young people to receive formal and informal support, to connect with each other, and to experience community. A public service participant described the need for “a space where youth can go…and connect with another youth who has perhaps gone through the system, is familiar with the system and to be able to join with them…” A young participant’s perspective supported this view.

> “Something you could benefit a lot from is just hav[ing] a safe place to to—having a safe place…somebody that they can actually like, confide in and …like, say whatever they need to, just having a …safe areas where they can …go and not be afraid of the place being torn down or being removed to put up something like condos.”

A similar idea was expressed by a participant who acknowledged the need for “a space to unload” amidst a community of people who “like, got your back,” whereas another young person missed the central gathering places for youth that an urban downtown could offer. Lack of easy transport to a gathering place for youth in the central city challenged youth people, and the absence of a “constant community,” as the same participant described, “is really one of the .biggest detriments to…youth mental health that I’ve seen or experienced just because I think it’s so important to …make those kinds of connections, and it really takes a lot of …initiative for kids in Seattle to do that.”

#### No one really knows how to help themselves

Youth participants reiterated that most young people lacked basic information about what to do if they experienced a problem with their mental health. School counseling services, in high schools and colleges, were thought to be insufficient for young people with significant needs and were often overwhelmed by demand. Several youth participants expressed the need for increased outreach and information about mental health care. One youth noted that school environments tend to be more reactive rather than proactive when communicating about mental health resources. “There isn’t very active promotion anywhere about mental health resources…word of mouth, I think is actually the biggest way of hearing about these resources…but like active encouragement and using them is not really present.”

#### Cultural barriers

Youth participants acknowledged the influence of cultural expectations on seeking help for mental health problems. One participant explained the implications of visibility when managing a mental health problem: “It’s very important that people know that if someone is dealing with the mental health issue, they’re not only dealing with their mental health, but they’re dealing with what people are going to say about them. They’re dealing with what their parents are going to think about them, what their grandparents are going to think about them.”

Young people believed that social stigma attached to mental illness was prevalent across most communities, but collectivist communities were also more likely to pull together to support each other. In some settings, such as schools, being associated with mental health groups or events could lead to stigmatization, causing young people to avoid them. One participant described “cultural stigma of mental health” as a deterrent to seeking care, particularly when mental health problems were not taken seriously. Relatedly, young people requested greater availability of culturally sensitive mental health services. They reported a dearth of non-white therapists and a missing “cultural connection” with therapists available to them. This included a need for providers who understood differing generational perspectives around mental illness as well as generational differences in exposure to mental health threats. A youth participant noted that young people compared their lives to their immigrant parents’ experiences and questioned, “…like, what right do I have to be sad about this?” A public service participant identified the lack of culturally appropriate community-based youth mental health support as a gap area, particularly as alternatives to more restrictive forms of care.

### You need to hear from the kids: youth leadership and youth participation

Youth/ally participants in our study were actively involved in promoting or leading projects, initiatives, or organizations interested in youth mental health. As such, they emphasized the importance of youth involvement in the development and promotion of youth mental health interventions or activities. For some community organizations, the value of youth participation was built into the structure, with one participant describing an organization where youth constituted 50% of the representatives on the board of directors. Others described youth-led exercises and projects where youth creativity, energy, humor, and words defined the intervention. They emphasized the importance of young people having the space and mandate to invent ways to relate to each other, and several observed “brilliant” and “poignant” activities resulting from these efforts. One participant, mused, “And when people see youth empowered in that kind of way, I think it really changes their perspective on their expectations of what these youth can do…in their life as well.”

Another participant described the level of youth leadership and involvement required to design shared spaces for youth mental health and a desire to see “without tokenizing…kids who have struggled with mental health care in Seattle in on the conversation surrounding mental health…they should be the ones working on the initiative…organizing employment and organizing what the [space] should look like…” Youth being integrally involved in the organization of the space was particularly significant because their pride in the project, i.e., their sense of ownership, would enable them to reach out to more young people, inviting them to join as organizers or co-creators of a shared space. Another participant emphasized the importance of designing interventions for specific communities, explaining that “mental health isn’t cookie cutter,” and ensuring a sense of belonging was a necessary part of design. This participant described the process of piloting a program, collecting feedback, and tailoring the program to meet the needs of participants.

Even with youth-led mental health activities, intentional outreach and a credible initiative were required for youth participation. Social media and word of mouth were identified as powerful and effective means for young people to inform others about mental health resources or programs. One participant discussed the challenge of credibility when disseminating information and encouraging participation in mental health activities, noting that the support of adult experts could lend credibility to young people. Partnering with people with subject matter expertise could safeguard against youths’ worries that they were leading conversations without the required knowledge.

On the other hand, young people also experienced their perceived lack of expertise by adults as a form of discrimination. Some participants spoke of youth input not being respected or their qualifications to talk about mental health being questioned. These beliefs played out in interactions in public spaces. “We would be at the table, but we would never get a chance to speak. We would never really get a chance to share our input. We’re kind of there to just be like a little model--to show off like, ‘look we have kids on our side.’” Others questioned more generally, “Do people actually want to hear our voice? Older people than us? Actually, want to hear our voice? Then do they actually act upon them?”

### Interventions

Challenges to youth mental health were most frequently attributed to social and relational problems, and the suggested interventions corresponded to these perceived causes. These ranged from creating positive and accepting environments that help “people feel wanted, involved, engaged, and valued” to implementation of specific services. Services that addressed family reconciliation after encounters with the criminal legal system or that worked to dismantle the “school-to-prison pipeline” were relevant and were viewed as part of efforts to address the harms of structural racism. Alternatives to therapy, like peer support, were also noted, as were more general approaches to stabilizing and supporting families.

Community institutions, such as schools, despite their challenges, were seen as important places to promote mental health. Youth leaders in the community actively engaged with schools by conducting workshops, mobilizing peers, and educating students, teachers, counselors, and educational personnel. They also explained how other community programs that engage young people in activities such as sports, theater, or shared hobbies could be used to explicitly introduce mental health themes or create communities that foster connection and support. For these programs to succeed, it was essential for young people to be centrally involved in their development and implementation, allowing them to contribute creatively. One participant remarked on witnessing young people create what they needed for mental health, “I think young people are so smart about making this stuff …joyful…Really, it was the most brilliant thing I’ve ever seen.” Youth participants articulated the nuance required for success: young people should not be tokenized in partnerships with experts or other adults, and they should also have access to people with knowledge and expertise they lack.

### Quantitative survey

One-hundred and seventeen participants completed the quantitative survey of intervention acceptability and feasibility (**Table 2**). Ninety-four (80%) identified as youth (e.g., under 35 years). Most (87, 74%) were women. Participants were broadly representative of the Seattle/King County population in terms of race: 57 (49%) were white, 18 (15%) were Asian, 11 (9%) were Southeast Asian, and 7 (6%) were Black American. Fifty (43%) were employed, and 65 (56%) were currently studying.

**S2 Table:**
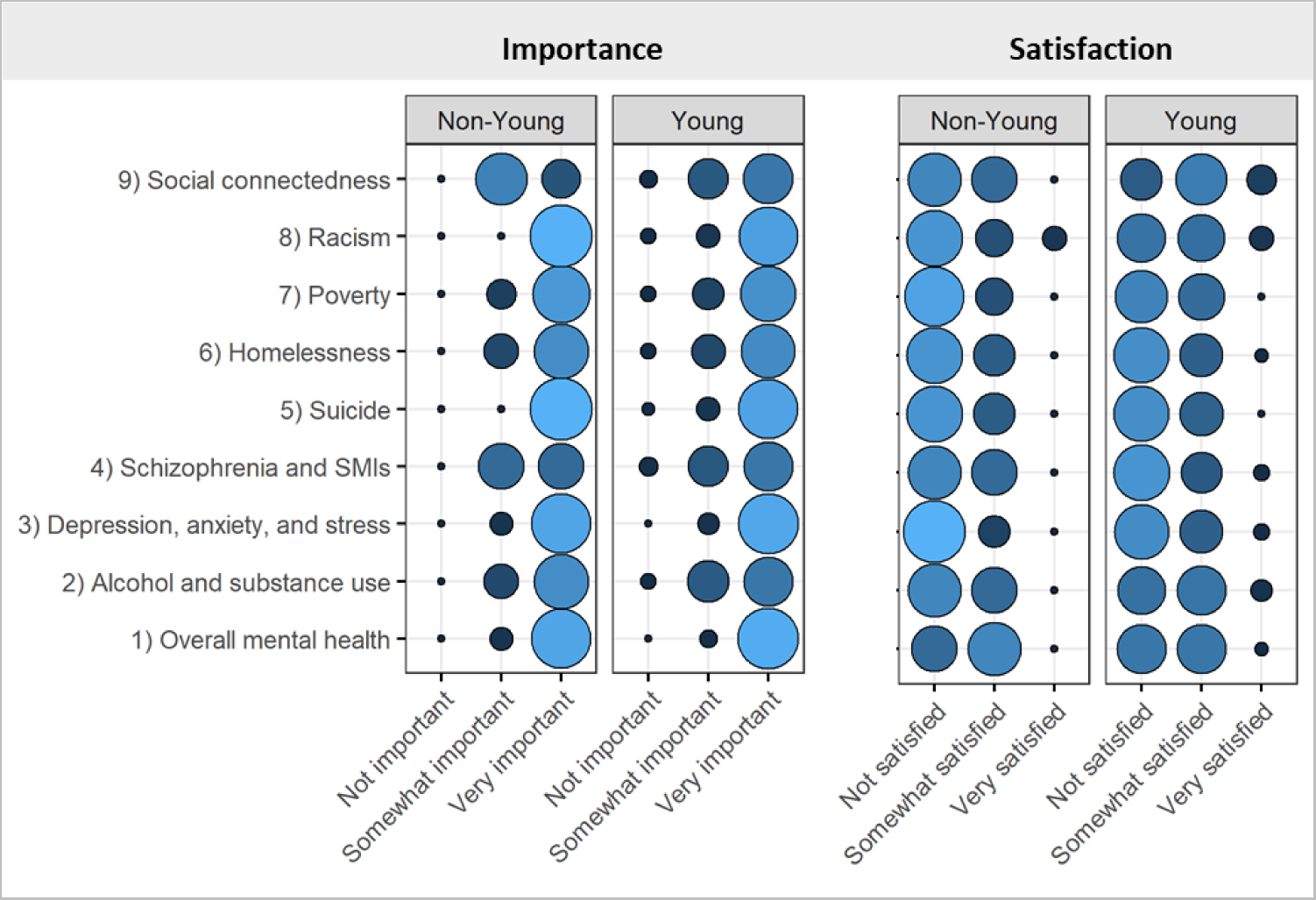
Questionnaire participant characteristics stratified by youth vs. non-youth (n=117)

**Figure 1** summarizes participant priorities regarding mental health, as well as their satisfaction with efforts to address those priorities. Almost all (97, 97%) endorsed that the overall mental health of young people in their community was ‘very important’. Similarly, almost all (94, 93%) endorsed the topic of depression as an important priority for young people in their community. There were notable differences between the perspectives of youth and non-youth participants. Youth participants placed a greater emphasis on the significance of social connectedness in their communities, whereas non-youth participants noted alcohol/substance use and suicide as problems in their communities. The satisfaction with efforts to address these issues was low across the board. However, youth participants were marginally more content with the efforts compared to non-youth participants. This distinction was most apparent concerning the promotion of social connectedness.

Young participants identified the lack of mental health awareness (48%), family support (46%), healthcare access (42%), and school support (39%) as the top barriers to positive mental health in their communities. On the other hand, non-youth participants identified the lack of economic opportunity (50%), mental health awareness, and the prevalence of violence (both 42%), as well as healthcare access, educational opportunity, and racism/discrimination (all 33%) as the top barriers.

Both youth and non-youth participants ranked schools as the highest priority location for mental health interventions (70%). Youth participants also mentioned online platforms (45%), community centers (22%), and health services (19%) as priority settings. Non-youth participants highlighted community centers (50%), online platforms (42%), as well as work and health services (both 8%) as priority settings. Churches/places of worship and public spaces were not commonly suggested as appropriate intervention platforms.

Both youth and non-youth participants ranked counselling and treatment as the highest priority service for young people (45%). Youth participants also identified other priority interventions such as resilience training (33%), positive messaging (32%), safe spaces (25%), awareness training (23%), housing and social services (21%), and employment (20%). In contrast, non-youth noted other priority interventions including positive messaging and housing/social services (both 33%).

Participants rated all four evidence-based interventions as highly acceptable (**Table 3**). Youth participants had a narrow preference for school-based mental health promotion, while non-youth participants rated both Friendship Bench and peer support services highly. There were no statistically significant differences in intervention acceptability between the two groups.

**S3 Table:**
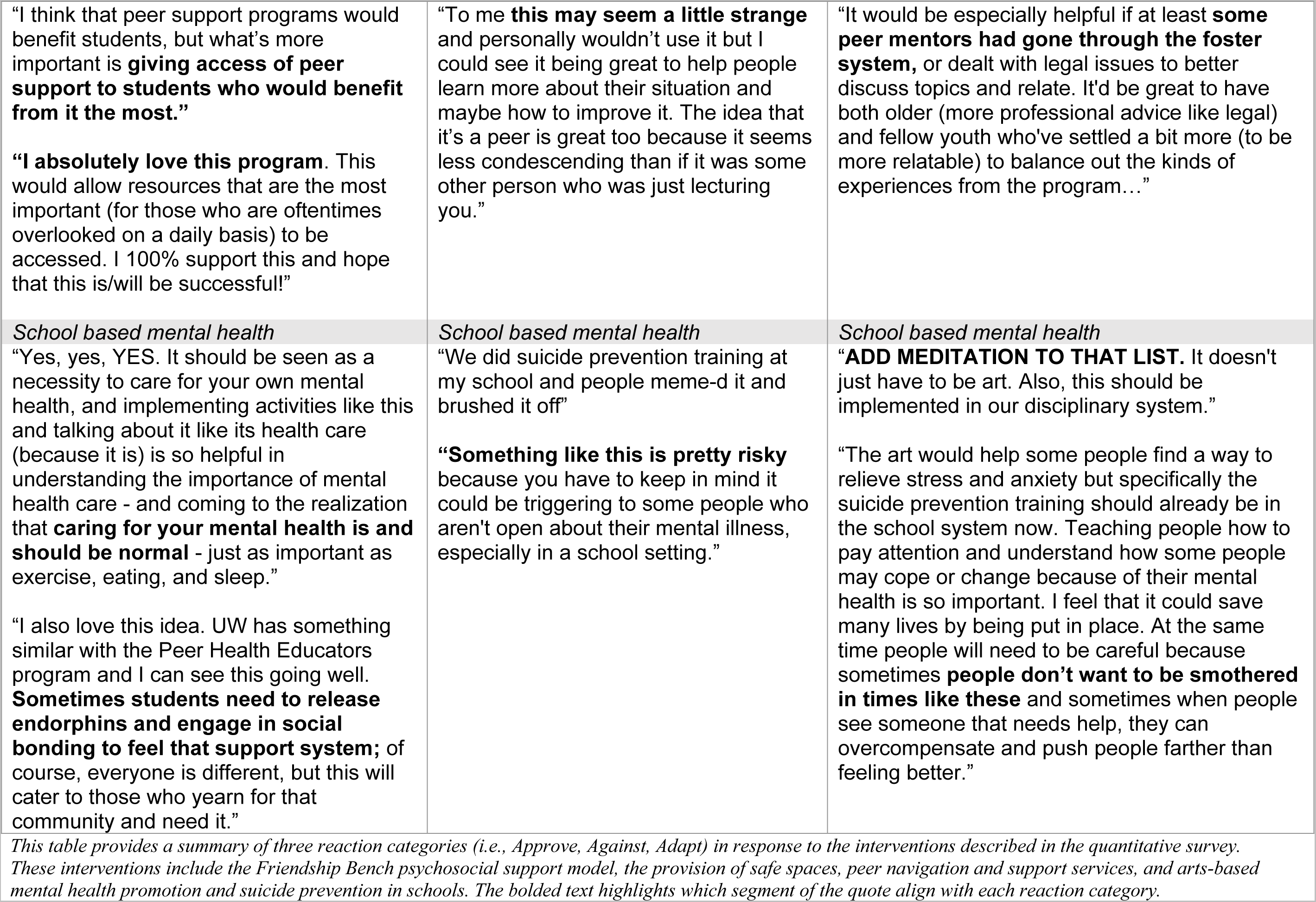
Intervention acceptability stratified by youth vs. non-youth (n=117)

### Open-ended feedback

Participants had the option to provide additional context to their positive or negative reactions, as well as options to adapt the interventions in the survey (**Table 4**). Suggested adaptations included adding a training component on social justice for individuals delivering psychological interventions (e.g., the Friendship bench); ensuring that spaces reserved for youth to gather were safe places for Black people as well as people of color; and encouraging the recruitment of people with lived experiences of adversities to participate in peer support programs.

**S4 Table:**
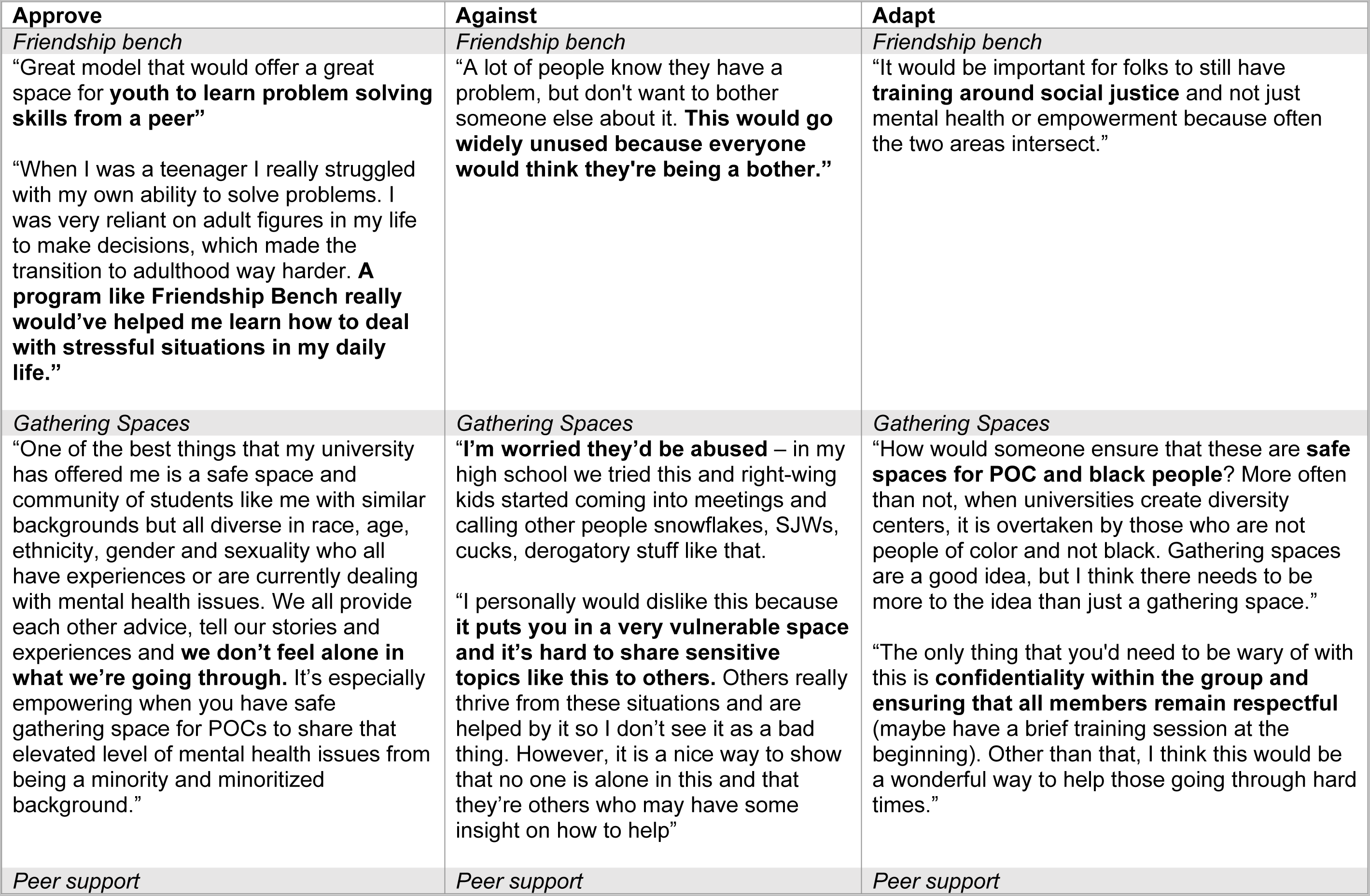
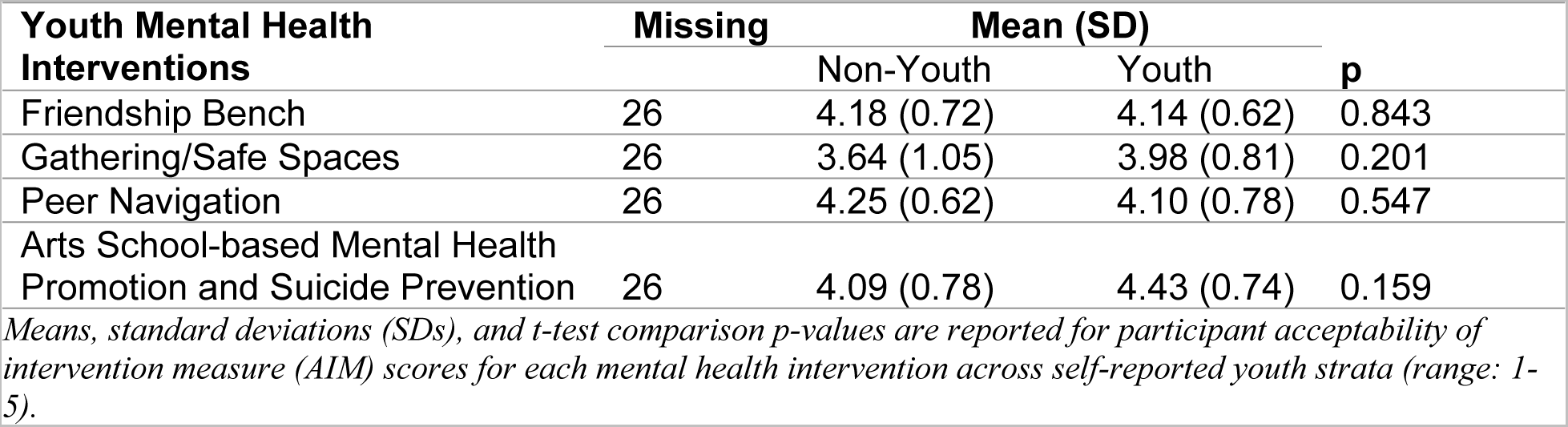
Open text comments on interventions described in the cross-sectional quantitative survey of youths and adults.

## Discussion

This study identified gaps in youth mental health services and opportunities for intervention and assessed the feasibility and acceptability of interventions that could support youth mental health in Seattle. Our qualitative key informants--young people/youth allies and public servants--emphasized unmet needs to reduce social isolation and lack of connectedness as key contributors to poor mental health among youth in Seattle. Quantitative survey data confirmed the need for greater access to mental health services and revealed differences in perceived acceptability of specific interventions between youth and non-youth participants. While both groups viewed evidence-based mental health interventions as highly acceptable, youth preferred interventions promoting social connectedness, peer support, and holistic approaches to care, while non-youth were more concerned with interventions focused on suicide, alcohol and substance abuse prevention. Youth participants also emphasized the significance of their active involvement in the development and implementation of mental health interventions, as it would improve relevance of the interventions to their needs. Given the regionally specific context of our study, generalizability to other populations may be limited. Nonetheless, our findings shed light on the concerns voiced by young individuals in Seattle, which align with those expressed by young people in other studies involving peer support, school-based interventions, and digital mental health interventions as well as the need for safe spaces for building connections in urban environments.

### Preferred Platform: Schools

Consistent with previous studies (Parikh et al. 2019; Arora and Algios 2019), participants in both the quantitative and qualitative studies identified schools as a priority setting for mental health interventions. School-based mental health programming has been effective in improving social skills, overall well-being, and reducing depressive and anxiety symptoms in some samples (22–25). Schools offer a built-in social community for young people and an opportunity to promote mental health awareness and provide counseling services. Connectedness to school is significantly associated with positive outcomes, including decreased risk of depressive and anxiety symptoms among young people (26). However, stigma is perceived as a significant barrier to accessing school-based mental health services, which was also emphasized by youth in our qualitative study. Nevertheless, schools provide a platform to involve students, teachers, school administrators, family and community members as potential stakeholders in the intervention design, as well as stigma reduction. In fact, “whole school” approaches that aim to integrate skill development into daily interactions and practices through collaborative efforts including all staff, teachers, families, and children have been found to improve the school ecosystem related to mental health (27–29).

### Preferred Platform: Digital

Young people are avid users of digital platforms in most parts of the world. In recent years, digital mental health interventions have become increasingly popular for early identification and treatment of mental health conditions given their utility for expanded access and youth-friendly nature. (30, 31). Our research found that young people in Seattle expressed a strong preference for online mental health platforms, which qualitative respondents emphasized as an important tool for engaging with and delivering mental health education, awareness, and potentially interventions. However, previous studies on youth preferences for online mental health platforms have yielded conflicting results, suggesting that different young people may have different needs and preferences (32). Some studies have found that young people prefer online mental health services due to the anonymity, privacy, and confidentiality they offer, as well as the reduced stigma associated with seeking care online (33–36). Other studies have found that young people prefer face-to-face services, or may not seek care at all (37).

### Preferred Mental Health Intervention: Psychological Interventions

Our quantitative analysis revealed that psychological interventions were the most preferred mental health interventions among young people. Youth participants rated the Friendship Bench problem-solving therapy intervention, which is currently being tested and implemented with adolescents in southern Africa (38, 39), as highly acceptable. Our key informants also stressed the need for improved access to standard mental health services that are culturally appropriate. Consistent with our findings, other studies have shown positive views of therapy among young people (40). While youth may be uncertain about their preferences for specific therapeutic approaches, they expressed a strong preference for meeting with a therapist alone, without parental involvement (41).

### Safe Space

Youth participants in our study emphasized the need for designated physical community gathering spaces and incentives such as free transit to increase access to safe public spaces. This aligns with prior research that highlights the effectiveness of safe and inclusive spaces in promoting youth mental health, social connections, and a sense of belonging. Establishing safe spaces in urban areas, such as downtown Seattle, could serve as a safe haven for many young people, promoting adolescent development, socialization, and mental health (42), as well as fostering social connectedness (43, 44).

### Peer-to-Peer Support

The availability of support from peers who share similar experiences was a key component of safe spaces in urban areas, as evidenced by the desire expressed by youth in our qualitative study for peer coaching, mentoring, and support. Peer-to-peer support can be integrated across various platforms, including schools, online venues, and safe spaces, and has the potential to address the barrier of stigma related to mental health identified in our study (45). While peer coaching for mental health has been studied in adults with mental illness, (46), fewer studies have focused on youth. However, online peer-to-peer support platforms for youth have been studied more extensively (47–51), and have shown positive mental health outcomes, including increased feelings of connectedness and reduced feelings of isolation. School-based peer-led mental health interventions have also been shown to be effective (52). Further research on peer coaching, mentoring, and support for youth mental health is necessary to determine its potential benefits for youth seeking mental health care and information.

### Upstream Mental Health Interventions

Improvement in youth mental health and well-being in Seattle will also require upstream interventions that promote structural and intersectoral responses and engage multiple stakeholders. Action steps from our roundtable discussion (**Table 1)** primarily focused on structural and upstream interventions, which merit further attention in Seattle, including how to implement such interventions and their feasibility. For instance, social and criminal justice actions such as reducing youth incarceration could change the trajectory of life for youth at risk for justice involvement and improve mental health in the long-term. A recent study on integrated youth services as an emerging model of care pointed to the need for flexible models of service delivery (53). Such interventions cannot solely rely on the feedback and input of mental health professionals, advocates, and policymakers, but also equally benefit from engaging young people. Consistent with our study, the researchers suggested co-designing interventions with the youth in the target population (53). The citiesRISE platform could play an instrumental role connecting young people and key stakeholders, establishing a systematic way to engage youth in shaping implementation of interventions in Seattle (17).

### Youth Engagement in Intervention Design

Young people in our study expressed a desire to be actively involved in the design, development, and implementation of mental health interventions, feeling excluded from the decision-making process. Researchers and mental health experts recognize the importance of engaging young people in discussions about their mental health and well-being as essential for their social and emotional development (54). A recent study found that young people aspire to be actively involved in their mental health initiatives but lack mental health literacy and awareness to participate fully (55). Policymakers and youth service providers can support meaningful participation by providing mental health literacy training and opportunities for raising awareness in schools and community settings. Personalizing interventions could significantly improve the nature of mental health care and the benefits provided to young people and families (56). Enabling young people as co-designers and incorporating their perspectives into mental health interventions could increase uptake by youth and improve mental health outcomes (56). Research and policy approaches can integrate youth perspectives using methods such as user-centered design (57, 58), community-based participatory research (59) and youth integration into policy settings (60). A recent Australian study on youth collaborative engagement in mental health intervention design highlighted the importance of building trusting relationships with key stakeholders and service providers to facilitate rapid policy change (61). Embracing youth-designed mental health interventions fosters shared participation and empowers young people to drive solutions that address their mental health needs.

### Limitations

There are several limitations to our study. First, our quantitative sample size was relatively small, with a total of 117 participants. As such, our findings may not be representative of broader views on youth mental health intervention feasibility and acceptability across Seattle. Participants in our quantitative study were recruited through mental health networks and digital platforms and were likely to have a particular interest in mental health, raising concerns of selection bias. Second, our quantitative assessment centered on a limited number of evidence-based interventions (EBIs) and did not include social and structural interventions that youth also valued. Despite the limitations, our quantitative sample captured the racial and ethnic diversity of Seattle, possibly enhancing the diversity of perspectives. Our mixed method approach and triangulation of data and contextual information (e.g., roundtable discussion) served as the primary efforts to address bias. The multistakeholder participants in this study displayed considerable agreement across findings that reinforced intervention directions for youth at greater risk of mental health disparities in an urban setting.

## Conclusion

This study highlights the need for interventions that support youth mental health in Seattle, particularly those that reduce social isolation and increase social connectedness. These problems have intensified during the years of the pandemic, and cities play a role in meeting these mental health needs. Schools and digital platforms were identified as preferred platforms for interventions, while psychological interventions and peer-to-peer support were the preferred mental health interventions. Engaging multiple sectors in the implementation of mental health interventions (e.g., education and health) and social interventions that can support mental health is a starting point.

Equally important, involving young people in the design and implementation of these interventions can improve their acceptability and uptake. Policymakers and mental health service providers should prioritize mental health literacy training and awareness-raising opportunities to support meaningful youth participation in mental health initiatives. Embracing youth-designed mental health interventions fosters shared participation and empowers young people to drive solutions that address their mental health needs.

## Data Availability

Data cannot be shared publicly because of privacy and confidentiality assurances included in the consent/assent documents for study participation. Given the sensitivity and regional specificity of some of the data, the participants did not consent to use of their data for additional research purposes.

## ACKNOWLEDGMENTS

We are grateful for the contributions of Dr. Meher Antia, through the University of Washington Population Health Initiative, to the roundtable gathering of key stakeholders in urban youth mental health. We thank Ms. Nissana Nov for her contributions to recruitment and data collection activities and Dr. Suresh Kumar for insightful comments on this manuscript.

## SUPPORTING INFORMATION

**S1 Figure 1:**
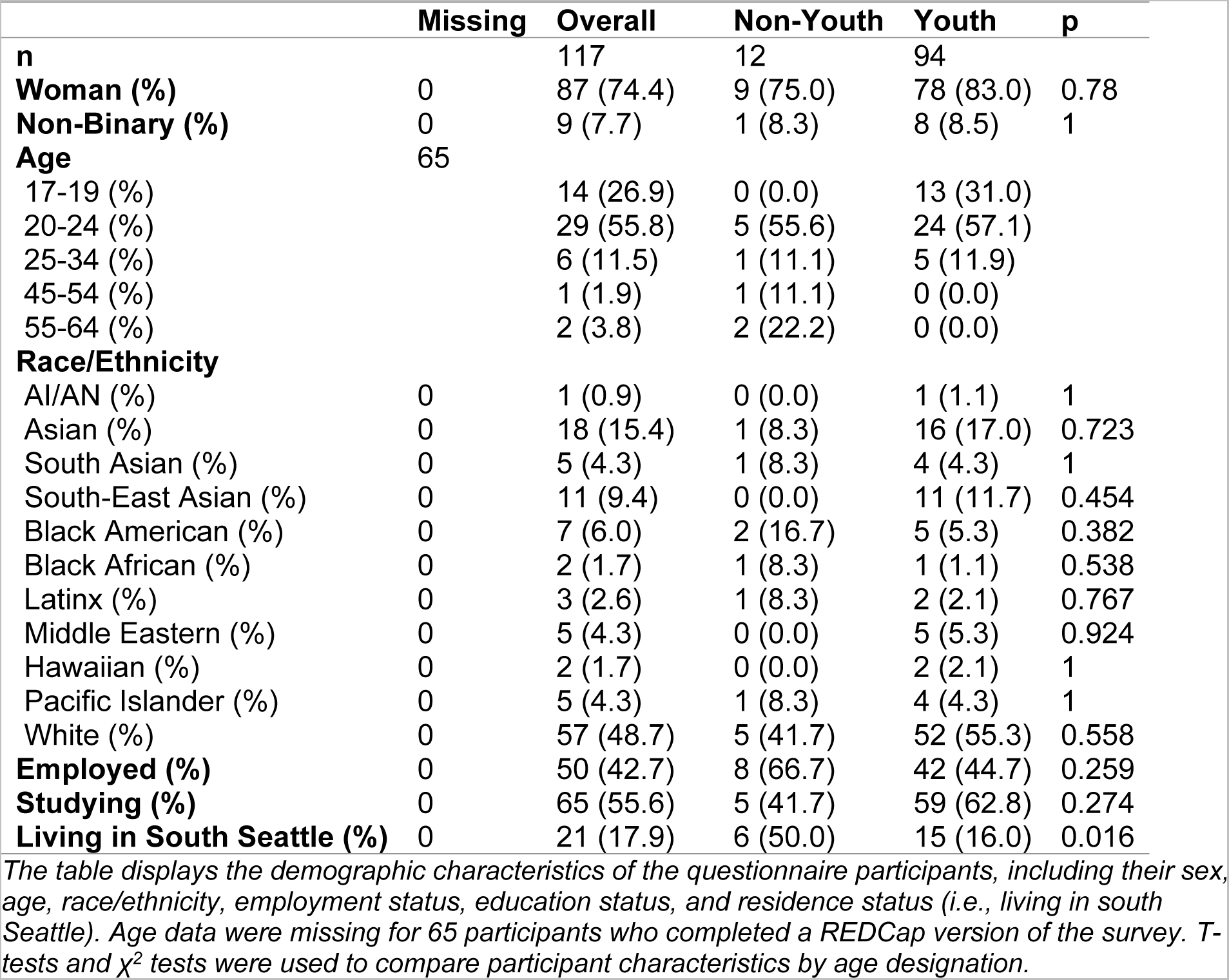
Priorities and unmet needs with regard to mental health (n=117. *This figure presents participant ratings of the importance of various mental health-related issues as well as ratings of satisfaction with current approaches to addressing those issues. Circle size and color correspond with the relative proportion of participant responses endorsing each importance or satisfaction level. Larger and lighter circles correspond with a greater proportion of participant endorsement. Responses are stratified by youth self-identification.*

